# Identifying Key Predictive Features for Opioid Use Disorder Using Machine Learning

**DOI:** 10.1101/2025.07.12.25331446

**Authors:** Suraiya Akhter, John H. Miller

## Abstract

**Background:** Opioid Use Disorder (OUD) continues to pose a pressing public health challenge across the United States, highlighting the critical need for early and accurate risk assessment tools that facilitate prompt prevention and intervention efforts. Machine learning methods have emerged as valuable tools for parsing complex medical datasets and aiding in clinical decisions. However, their effectiveness and interpretability largely rely on the appropriateness and quality of selected input features.

**Objective:** In this work, we conducted a comprehensive comparison of three distinct feature selection strategies—Alternating Decision Tree (ADT)-based scoring, Cross-Validated Feature Evaluation (CVFE), and Hypergraph-Based Feature Evaluation (HFE)— to identify the most predictive indicators of OUD.

**Methods:** The analysis was performed using data from the 2023 National Survey on Drug Use and Health (NSDUH), a dataset compiled by RTI International under the direction of the Substance Abuse and Mental Health Services Administration (SAMHSA). This dataset encompasses a broad spectrum of features related to demographics, behavior, mental health, and substance usage. Each feature selection method yielded a set of important predictors, which were subsequently used to train eXtreme Gradient Boosting (XGBoost) classification models. To enhance model transparency and interpretability, SHapley Additive exPlanations (SHAP) was employed to illustrate the influence of individual variables on model predictions.

**Results:** The performance of the models was evaluated and compared, with the model informed by CVFE-selected features achieving the best outcomes—demonstrating a predictive accuracy of 79.11% and an area under the curve (AUC) of 0.8652. The top 10 most influential features, based on SHAP value rankings from the best-performing model, included past-year misuse of pain relievers, recent alcohol use disorder, age group, history of asthma, receipt of substance use treatment in the past year, educational attainment, household size, total household income, marital status, and race/ethnicity. The web application, accessible via https://shiny.tricities.wsu.edu/oud-prediction/, offers prediction outcomes, probability metrics, and a SHAP visualization generated from the best model built using cross-validation-based approach.

**Conclusions:** The findings highlight the crucial importance of effective feature selection in enhancing both model accuracy and interpretability, ultimately supporting the development of practical, data-driven approaches that may help healthcare providers assess OUD risk and tailor prevention strategies to individual needs.

**Trial registration:** Not applicable as this research is not a clinical trial.

## Introduction

Since the early 2000s, the United States has experienced a dramatic escalation in both opioid use disorder (OUD) and overdose-related fatalities, culminating in the declaration of a nationwide public health emergency in 2017 [1, 2]. In that year alone, over 70,000 drug overdose deaths occurred, with a significant rise in cases involving fentanyl and its analogs [3]. Although opioid prescription rates have steadily declined since 2012, many individuals continue to receive long-term opioid treatments, which are strongly linked to heightened OUD risk [4–6]. In primary care, a substantial proportion of patients on chronic opioid therapy meet diagnostic criteria for OUD [5], and a nontrivial percentage of pain patients develop prescription-related OUD [6]. In the United States, millions are affected by OUD across both prescription and non-prescription opioid use [7, 8].

Traditional tools for identifying OUD risk—such as screening questionnaires and urine drug screens (UDS)—have important limitations. UDS can fail to detect drug use, especially when not directly observed, and self-reports are often inaccurate [9–11]. Consequently, many patients with OUD remain undiagnosed, leading to elevated risks for overdose and poor outcomes. Despite weak evidence supporting the long-term benefits of opioid therapy, healthcare providers frequently continue prescribing opioids for chronic pain conditions [12]. However, accurately identifying individuals vulnerable to OUD before starting opioid treatment remains a significant challenge. Established tools like the Opioid Risk Tool (ORT) and SOAPP-R have shown only limited utility [13]. Regression-based studies have attempted to identify predictive factors [14–19], but these approaches often struggle with issues such as class imbalance and poor generalizability [20, 21].

The application of machine learning techniques has gained considerable attention in the prediction of OUD, largely due to their ability to analyze and interpret high-dimensional datasets derived from electronic health records. These algorithms have been increasingly utilized to identify individuals at elevated risk by drawing from diverse data environments, including clinical records and commercial data repositories, and by modeling patterns across medical history, healthcare utilization, and behavioral attributes [3, 22–27]. A broad array of factors—such as gender, racial or ethnic background, educational attainment, household income, marital and employment status, geographic context, history of substance use, psychiatric conditions, prior engagement with treatment services, chronic diseases, criminal justice interactions, and type of health insurance—can all influence patient outcomes. However, the intricate relationships among demographic, behavioral, and clinical variables continue to pose challenges for reliable risk differentiation. To date, limited research has systematically assessed the significance or comparative influence of these individual factors. There is a clear demand for advanced machine learning models capable of evaluating feature importance across heterogeneous population groups.

We hypothesize that a machine learning framework can be effectively used to autonomously discern and prioritize critical variables for accurate risk estimation, thereby enabling more customized and data-informed approaches to care. Such models could inform tailored treatment decisions and improve care for patients with—or at risk of—OUD and associated comorbidities. To address this, we used data from the 2023 National Survey on Drug Use and Health (NSDUH), conducted by RTI International for the Substance Abuse and Mental Health Services Administration (SAMHSA). We built a foundational machine learning pipeline aimed at forecasting OUD risk, with a strong emphasis on comprehensive feature selection methodologies. By systematically investigating both the individual impact of predictor variables, our goal is to optimize the relevance of clinical and behavioral information in guiding effective, person-centered care. This work aspires to contribute to the growing field of precision medicine and reinforce the role of machine learning in addressing public health challenges. Accessible at https://shiny.tricities.wsu.edu/oud-prediction/, the web-based platform delivers a robust prediction framework that incorporates the cross-validation driven feature selection method. It provides SHAP-based explanations, supports batch processing of multiple samples, and allows integration of additional data to continuously enhance the model’s predictive accuracy.

## Materials and methods

An overview of the proposed methodology is depicted in Figure 1. The workflow begins with the collection of data for two groups—individuals diagnosed with opioid use disorder (positive cases) and those without the condition (negative cases). From this data, a wide range of potential features is generated. Feature evaluation techniques are then applied, including Alternating Decision Tree (ADT) [28, 29], Cross-Validated Feature Elimination (CVFE) [30], and hypergraph-based feature evaluation (HFE) [31], to systematically eliminate features that contribute minimal or no predictive value. The refined feature subsets are subsequently utilized to train a machine learning algorithm, enabling the evaluation of model performance in predicting OUD outcomes.

**Figure 1.**
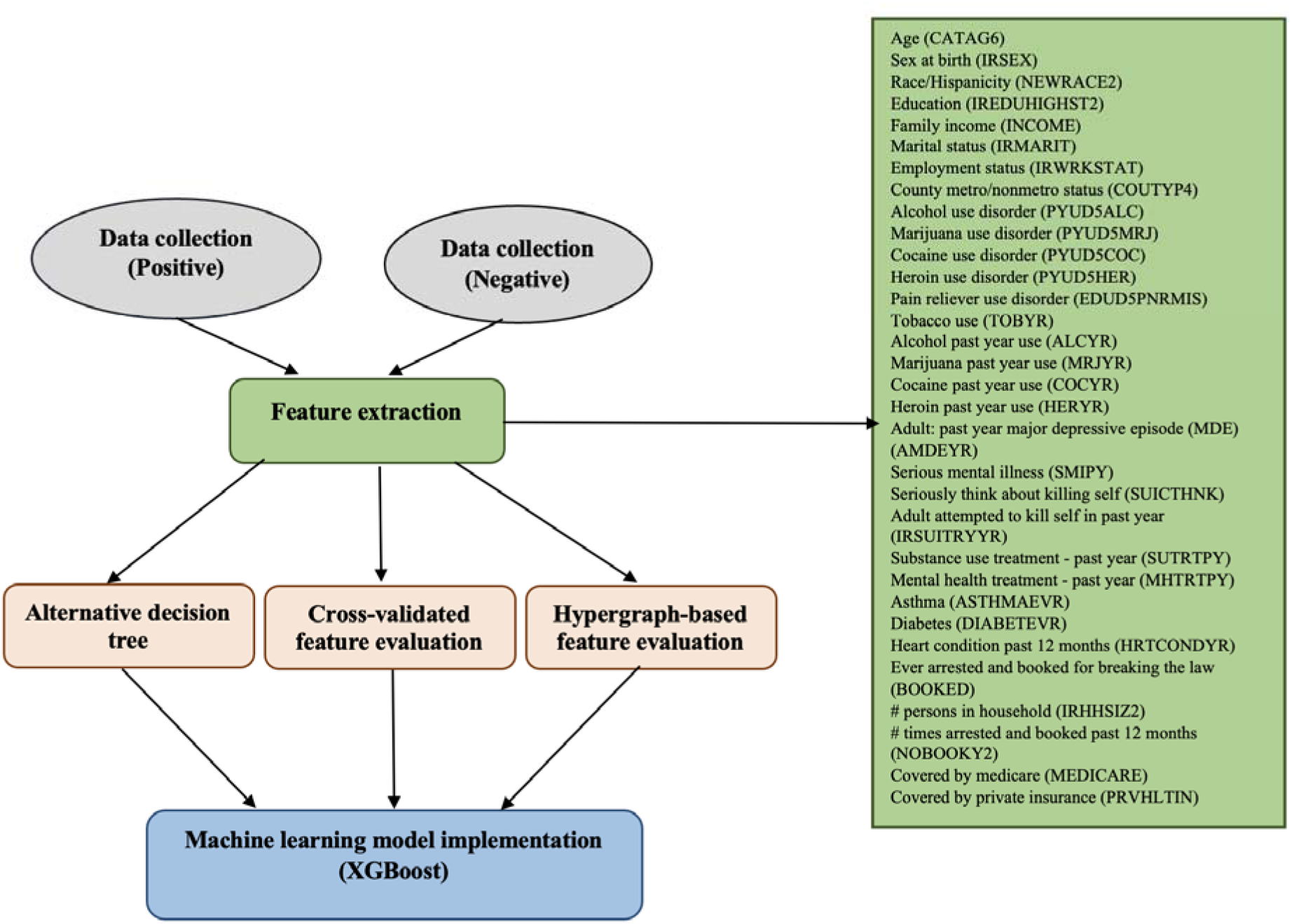
Workflow illustrating the procedure for opioid use disorder detection.

### Study population and features

This study employed data from the 2023 edition of the National Survey on Drug Use and Health (NSDUH), administered by RTI International on behalf of the Substance Abuse and Mental Health Services Administration (SAMHSA) [32]. The dataset comprises an extensive set of variables covering demographic details, behavioral patterns, mental health indicators, and substance use information. The final dataset included 791 participants diagnosed with opioid use disorder and 42,607 participants without the condition. To mitigate the issue of class imbalance, random undersampling was applied to the non-OUD group, reducing it to 791 cases and achieving a balanced dataset across both classes. To build the predictive model, 80% of the dataset was allocated for training purposes, and the remaining 20% was set aside for performance evaluation. A detailed breakdown of the variables, their categorical groupings, associated demographic distributions, and statistical significance values is presented in Table 1. Features exhibiting a *p*-value below 0.05 were regarded as statistically significant indicators. The target outcome—opioid use disorder—was defined based on the variable UD5OPIANY (Opioid Use Disorder - Past Year Users), where a value of 1 (Yes) indicated classification as an OUD case. In total, 32 features were shortlisted for analysis, all of which are listed in Supplementary Table S1 **(Supplementary Material)**.

**Table 1.**
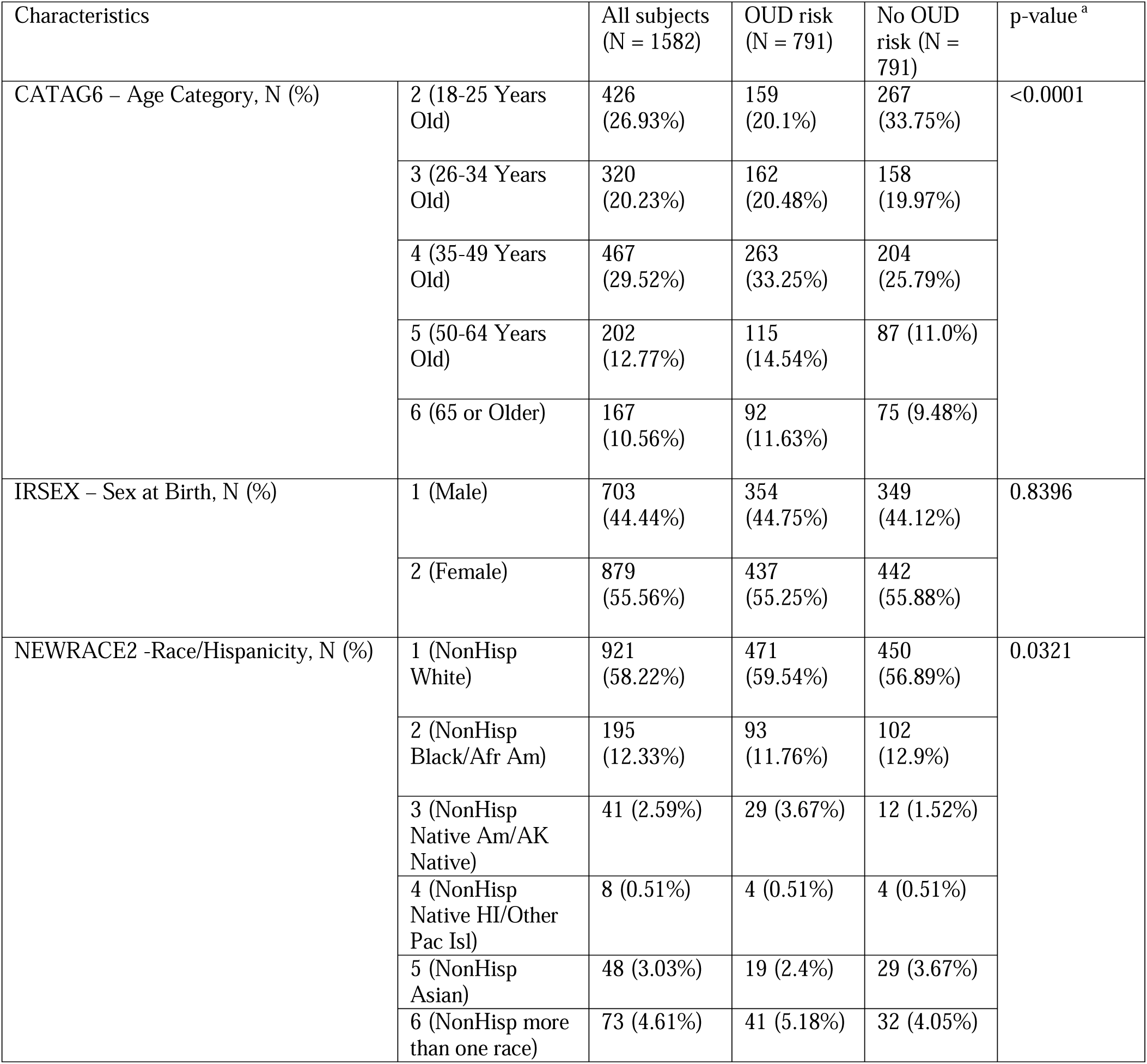
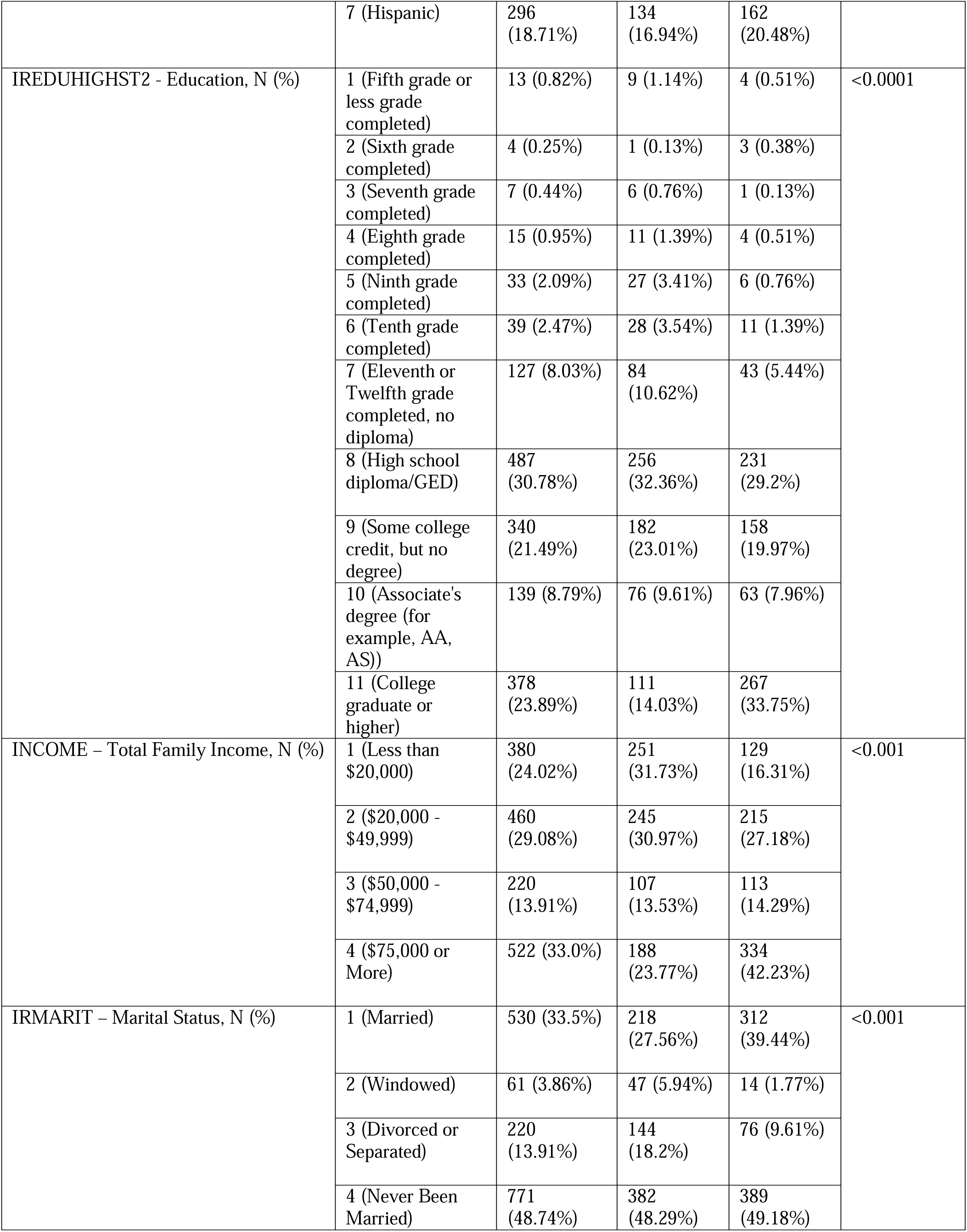
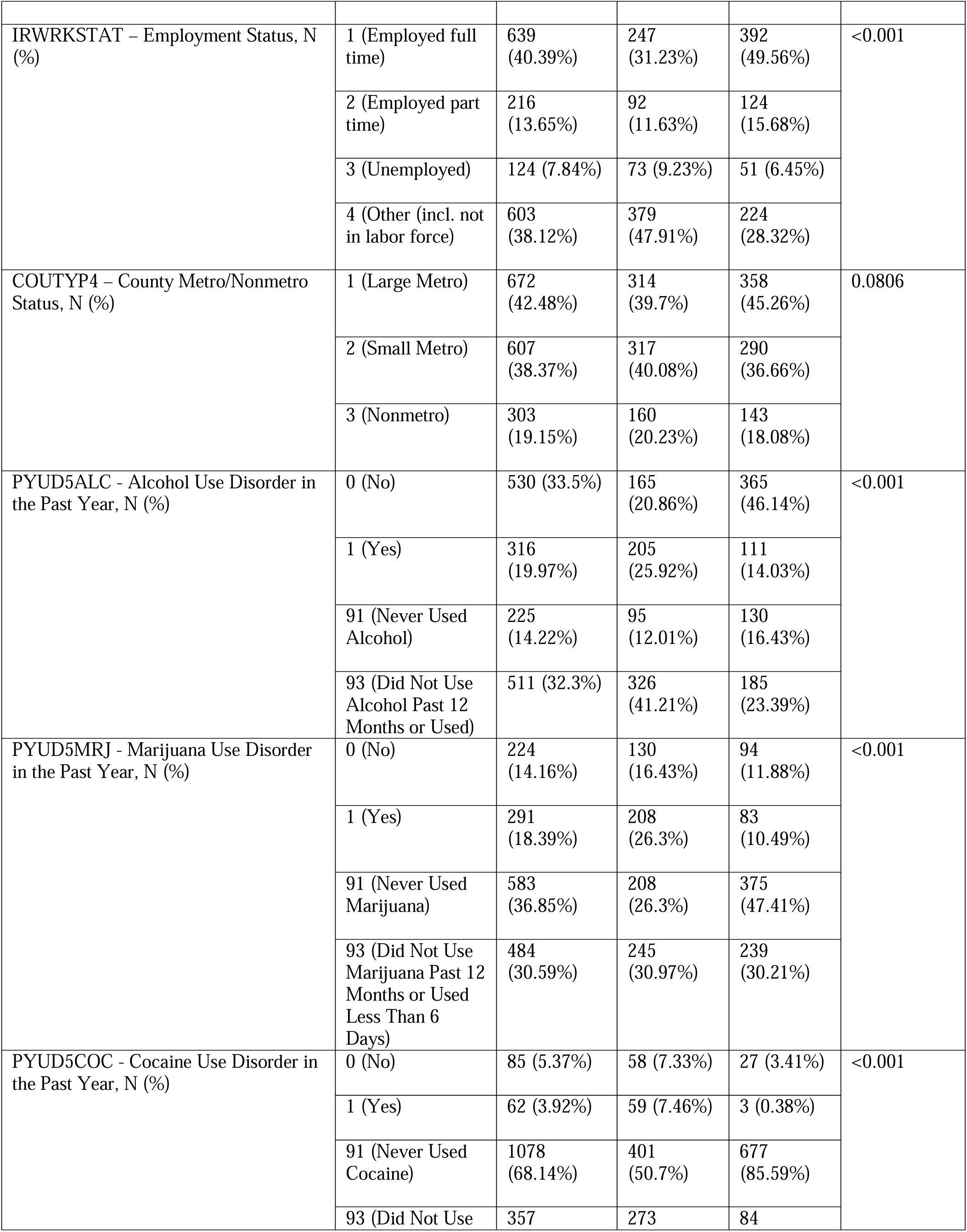
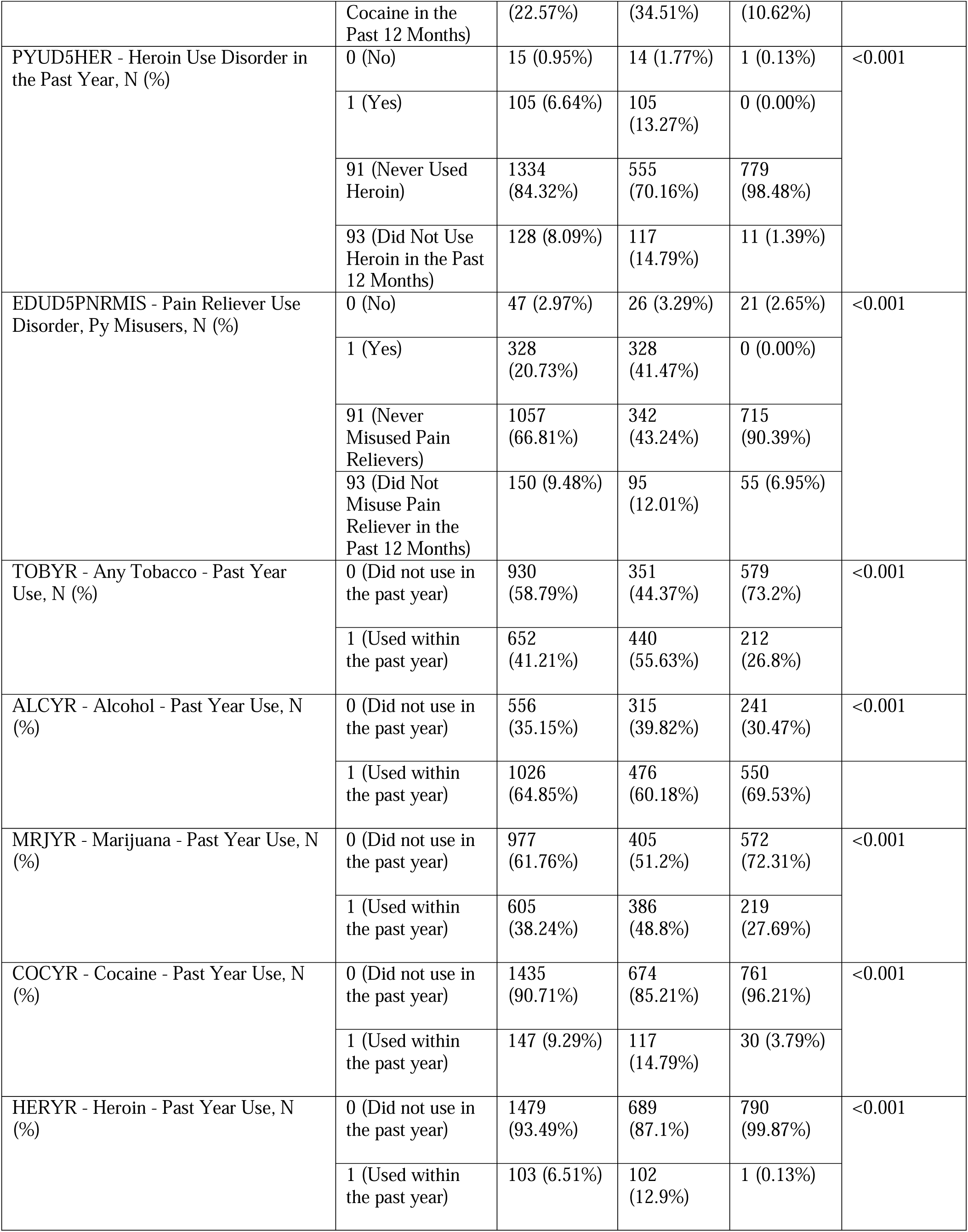
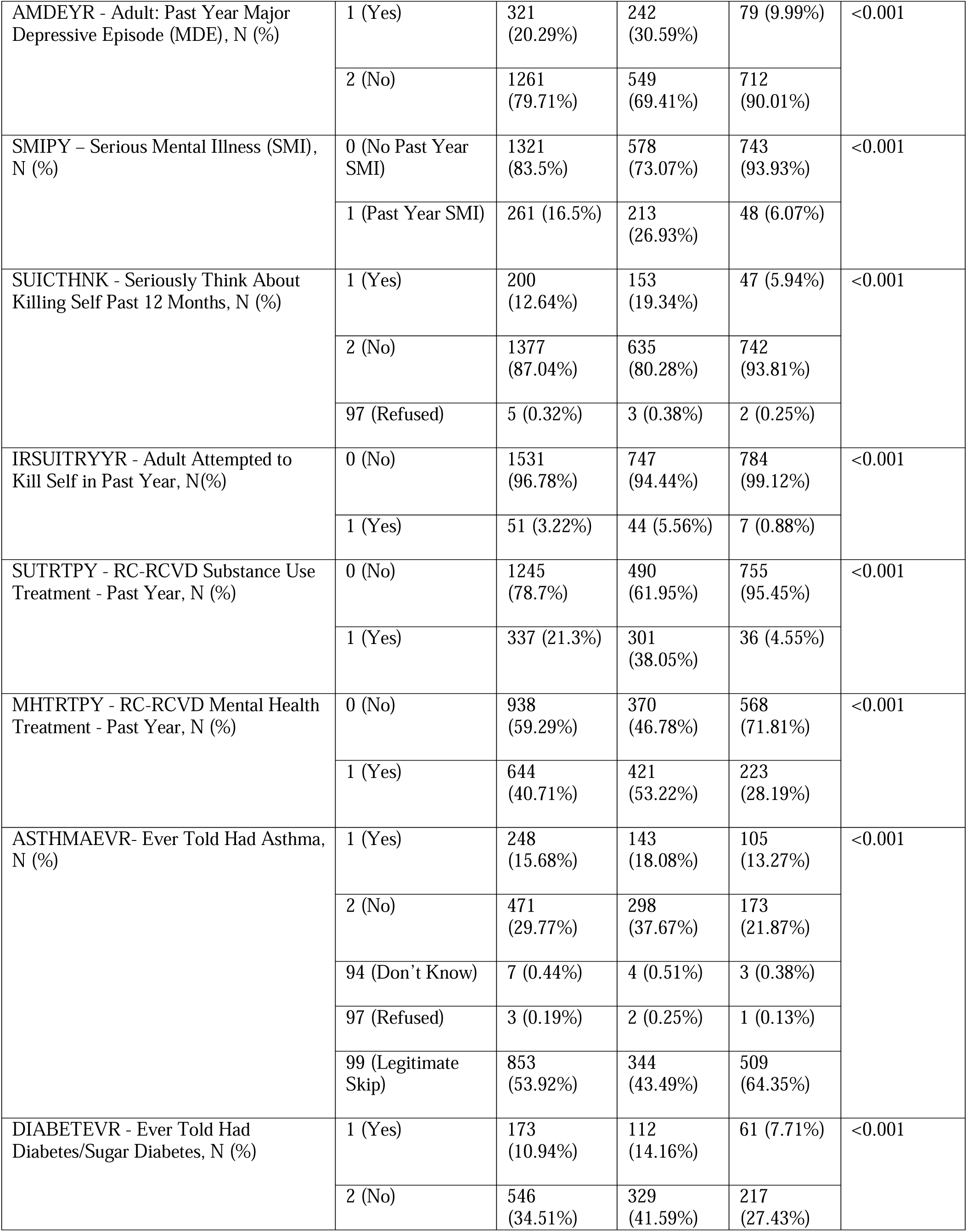
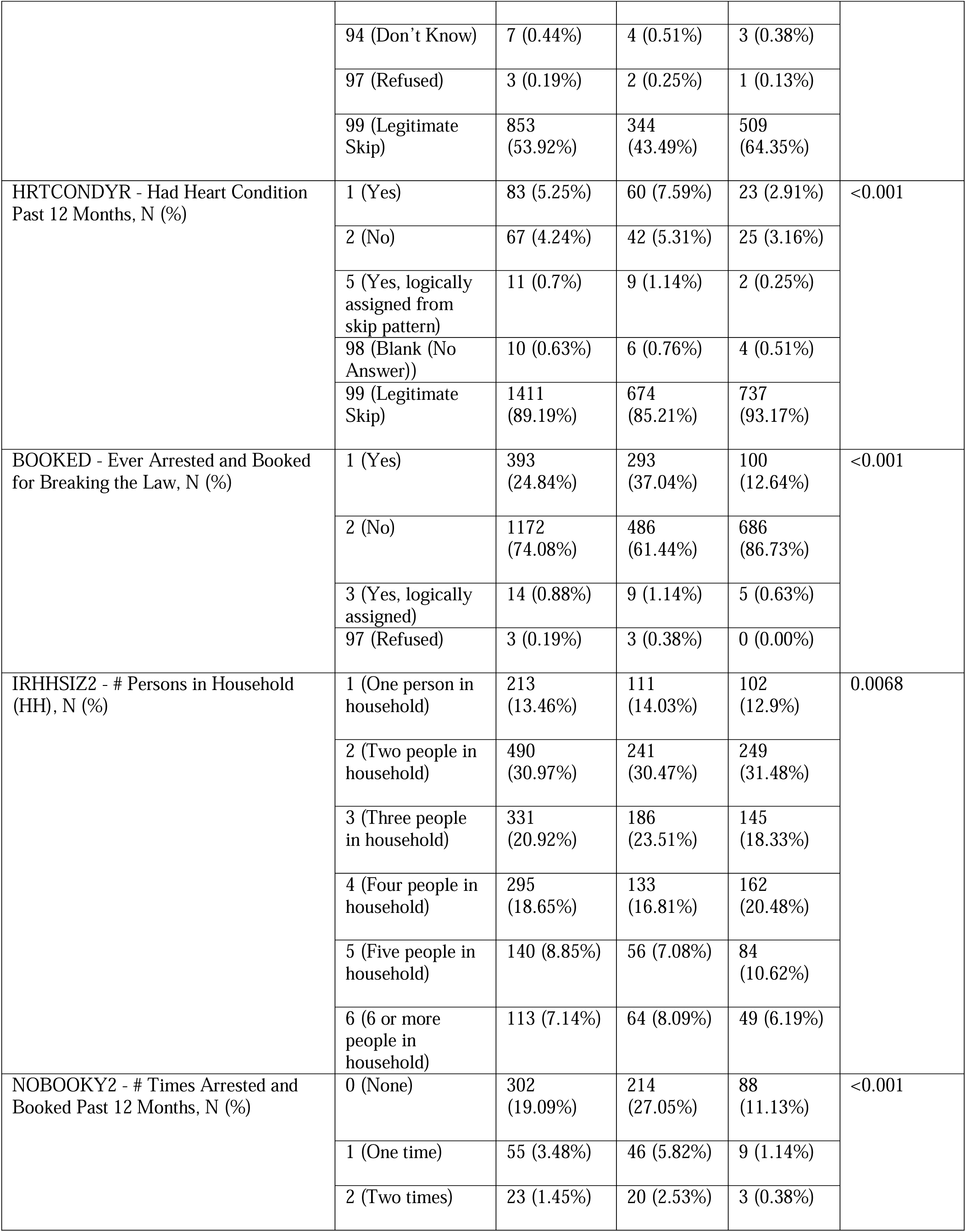
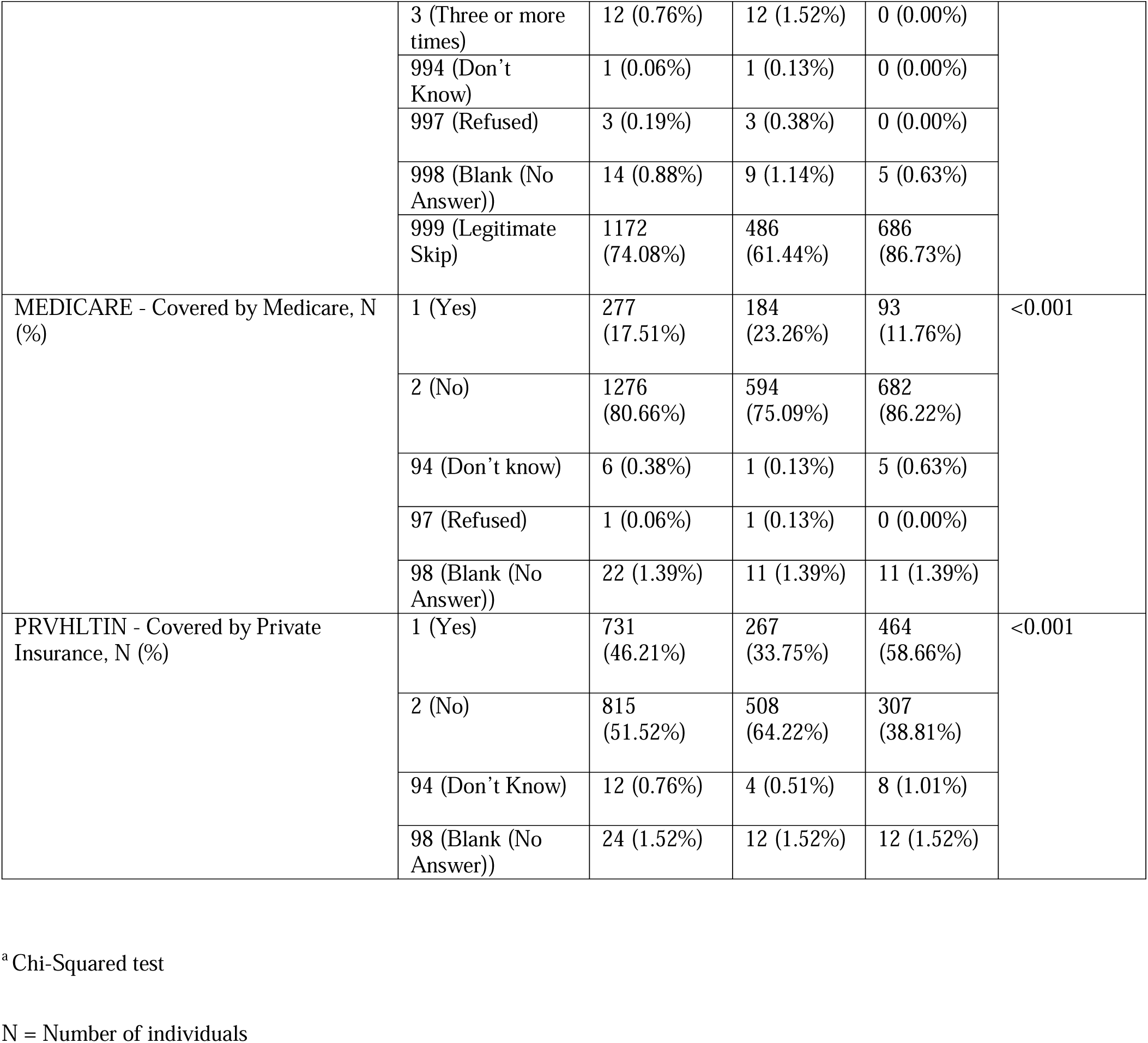
Overview of participants’ demographic and clinical profiles.

### Evaluation of features

Prior to model development, removing irrelevant or low-importance features is critical to enhance predictive accuracy. In this study, we applied three distinct feature selection methods—Alternating Decision Tree (ADT) [28, 29], Cross-Validated Feature Elimination (CVFE) [30], and Hypergraph-based Feature Evaluation (HFE) [31]—to systematically discard less informative variables. The ADT algorithm combines the straightforward interpretability of a traditional decision tree with the superior prediction capability afforded by boosting methods. It encodes knowledge as a tree structure composed of interconnected decision stumps, a technique often employed in boosting frameworks. Unlike conventional trees where branches are mutually exclusive, ADTs allow overlapping branches, enabling more flexible decision paths. The model’s root is a prediction node that assigns a numerical score, followed by layers of decision nodes representing predicate conditions or tests. These layers alternate between prediction and decision nodes throughout the tree. Importantly, prediction nodes appear both as the root and as terminal nodes (leaves), distinguishing ADTs from classic decision trees and highlighting their unique architecture and operational logic.

The ADT constructs a collection of rules, each comprising a prerequisite, a condition, and two corresponding scores. Conditions take the form of predicates structured as “attribute <COMPARISON= value,” while prerequisites are logical conjunctions of such conditions. Rule evaluation proceeds through nested conditional statements, using the associated scores to generate predictions for individual data points. The process begins with a root rule, whose prerequisite and condition are both set to “true,” and whose scores are computed based on the weighted distribution of training samples. Initially, all training instances are assigned equal weights, calculated as the reciprocal of the total number of samples. The algorithm then proceeds iteratively, generating new rules by selecting the optimal pairing of prerequisites and conditions that minimize a specific objective function, denoted as *z*. This function quantifies how effectively a rule separates positive from negative instances, guiding the search for the best rule to add. At each step, the scores for the newly created rule are updated using a boosting strategy, and the weights of training samples are adjusted in accordance with the accuracy of the new rule’s predictions. The training cycle continues until a stopping criterion is met, which may be a predetermined maximum number of iterations or a threshold for minimal accuracy improvement. The ensemble of rules produced by the ADT forms an alternating decision tree, characterized by prediction nodes containing numeric values, and structured according to the logical prerequisites defined by each successive rule. Notably, the ADT model employs only a subset of the total available features in its construction. In our implementation, we evaluated 50 randomly chosen values for the boosting iteration parameter, *B*. The resulting ADT-derived decision tree is displayed in Supplementary Figure S1, highlighting the subset of 32 candidate features selected. This process yielded a final set of 10 features, detailed in Supplementary Table S2 (**Supplementary Material**).

In addition, we incorporated the CVFE approach together with a hypergraph-based technique to refine the initial pool of features. The step-by-step process of the CVFE method is illustrated in Figure 2. Initially, the dataset was randomly divided into *c* distinct subsets. For each subset, the XGBoost algorithm was applied to determine the most influential features, with model hyperparameters optimized through a grid search strategy. Feature selection was conducted separately on every subset, after which an intersected feature set—comprising features common across all subsets—was identified. This entire procedure was repeated *e* times, generating *e* intersected feature sets in total. To finalize the feature selection, any feature appearing in at least (*p* × 100)% of these intersected sets was included in the ultimate chosen feature set. Table 2 presents the count of features identified in these subsets across various configurations of the parameters *c*, *e*, and *p*. The detailed lists of these key features can be found in Supplementary Tables S3-S7, located in the **Supplementary Material**.

**Figure 2.**
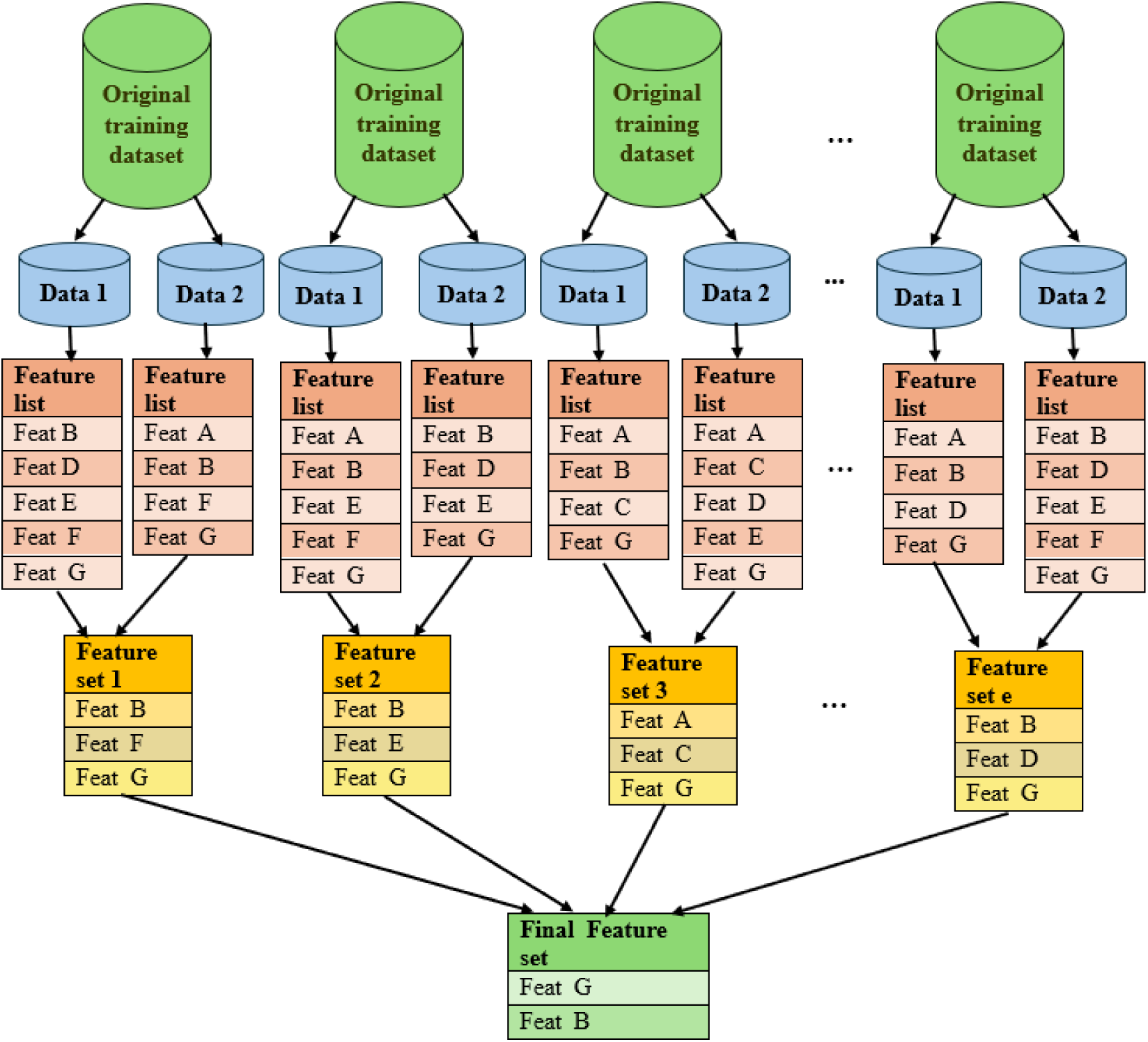
llustration of the feature selection process employing the cross-validated feature evaluation method.

**Table 2.**
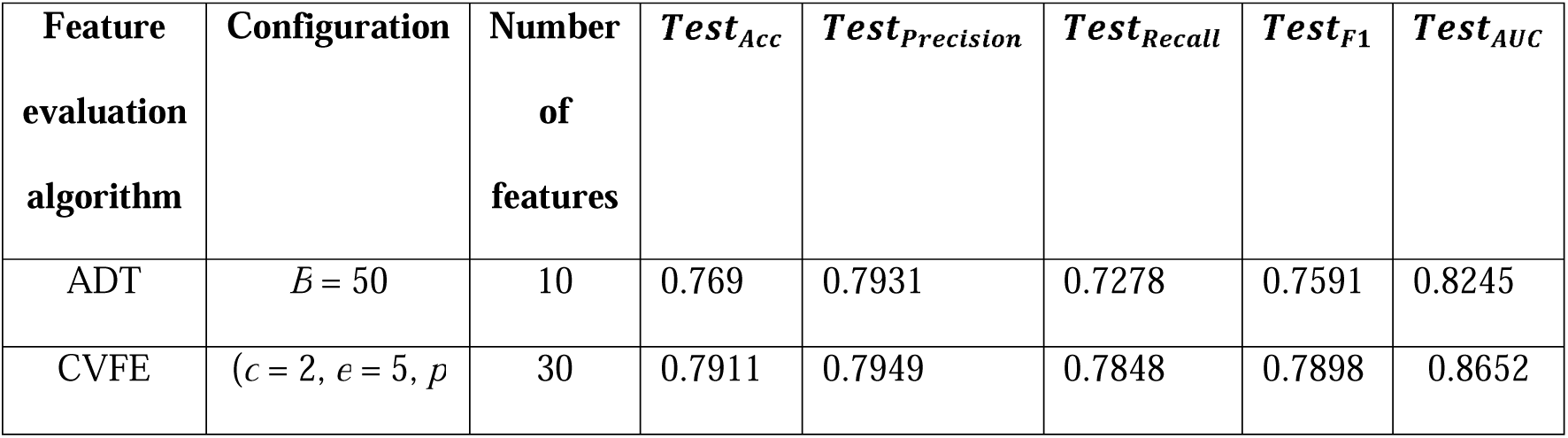
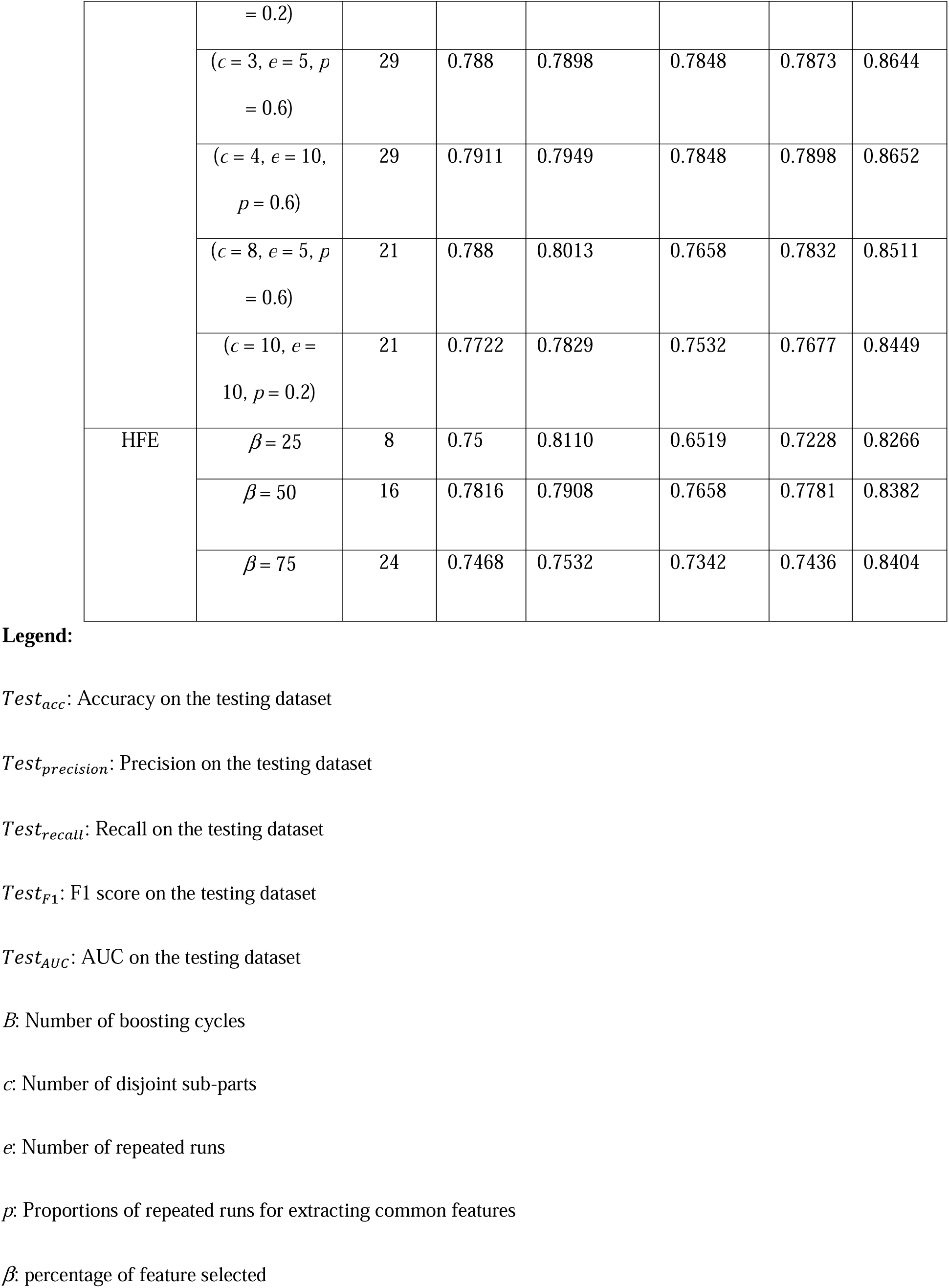
Performance evaluation on the test set, detailing the count of selected features and associated metrics: accuracy, precision, recall, F1 score, and AUC for each feature selection method.

The HFE technique involves generating a hypergraph structure from the set of input features. Unlike standard graphs, which are denoted as G(*V*, *E*) with vertices (*V*) connected by binary edges (*E*), a hypergraph generalizes this concept by permitting each edge—referred to as a hyperedge—to link multiple vertices simultaneously. Formally, a hypergraph is represented as G(*V*, *E*), where *V* is the set of nodes and E comprises the hyperedges, each of which is a subset of *V*. Figure 3 provides a visual comparison between a traditional graph and a hypergraph, while Supplementary Figure S2 (**Supplemetary Material**) presents the hypergraph structure derived from the full training dataset used in this study. In the HFE framework, feature relevance is determined by assigning weights to the hyperedges associated with specific feature values. These importance scores are computed using the principle of hypergraph cut conductance minimization [31], resulting in a feature rating vector that reflects the relative influence of each feature. The scores in this vector are ranked, and the top *z* features are selected. The value of *z* is defined as β × *m*, where *m* is the total number of features and β represents the desired proportion of features to retain. In our analysis, the resulting number of selected features for different values of β are listed in Table 2. Supplementary Tables S8-S10 (**Supplementary Material**) provide the specific feature subsets obtained for β values of 25, 50, and 75, respectively.

**Figure 3.**
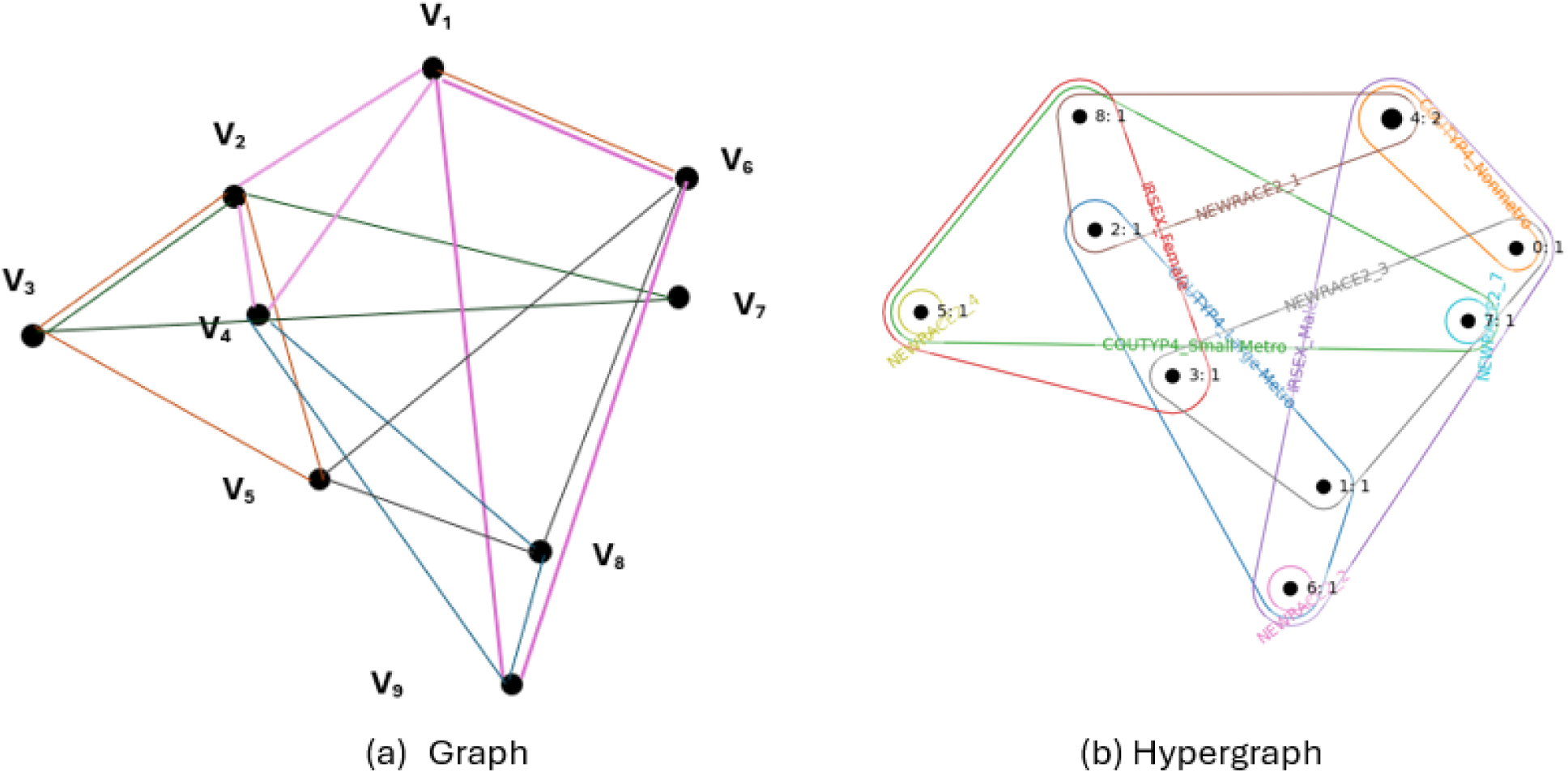
Visualization comparing the structural differences between a standard graph and a hypergraph.

### Web application

Figure 4 illustrates the integration of cross-validation-based feature assessment method within a machine learning-driven web application. This platform provides classification results along with probability estimates. The user interface guides users through dataset upload, binary classification execution, and probability calculation. Users can download output files and add new training samples to enhance the model’s accuracy. Additionally, the application enables exporting a SHAP plot to explore the impact of key features on predictions. The web application can be accessed publicly at https://shiny.tricities.wsu.edu/oud-prediction/.

**Figure 4.**
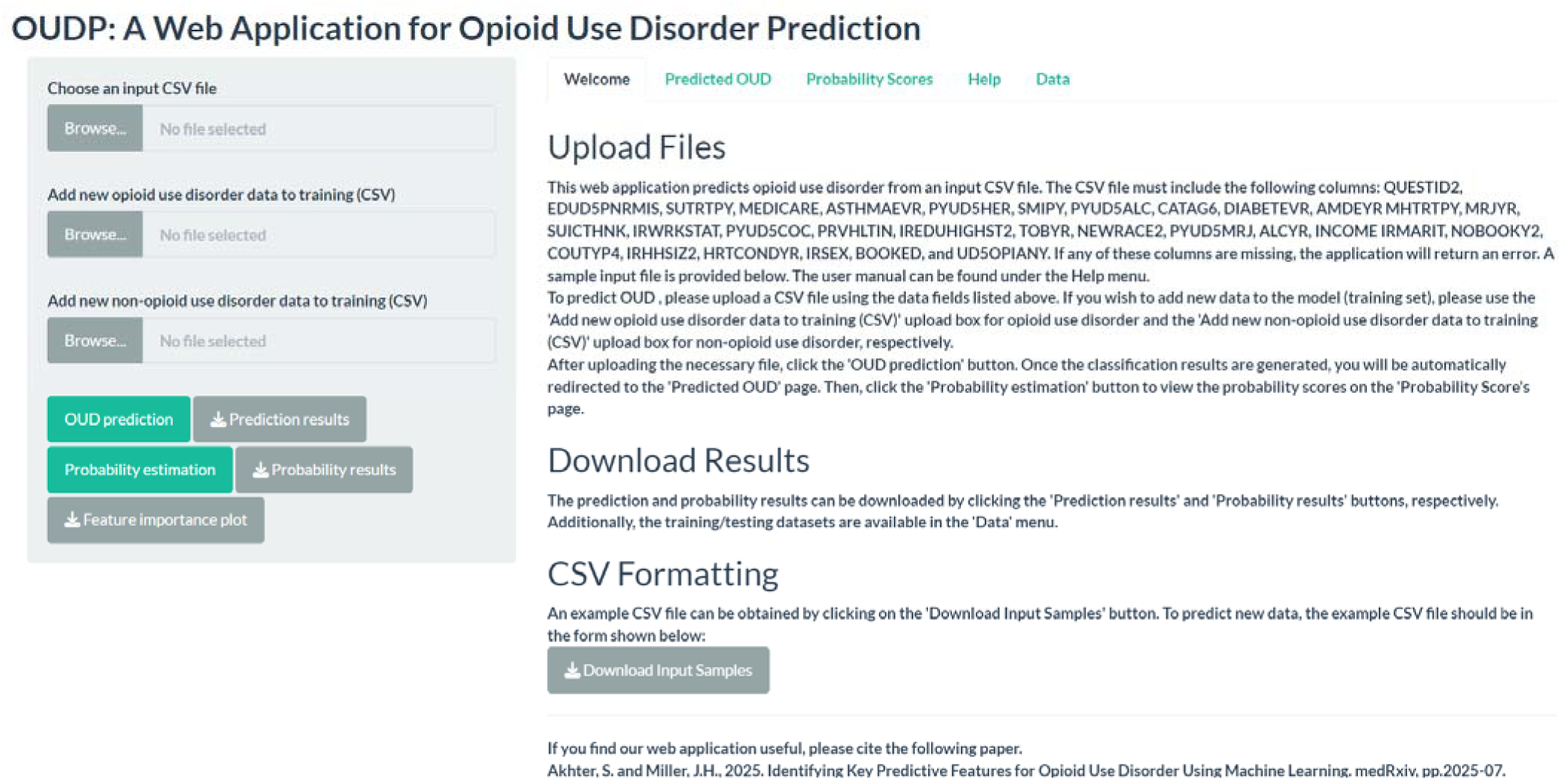
The web application for opioid use disorder prediction.

### Code and data availability

All experimental datasets and the corresponding code are available in the repository at https://github.com/suraiya14/OUDP, as also referenced in the **Supplementary Material**.

## Results

Following the reduction of the original feature set using three independent feature selection strategies, we developed individual predictive models based on the refined subsets, utilizing the XGBoost algorithm [33]. To interpret and quantify the impact of each feature on model outcomes, we applied the Shapley Additive Explanations (SHAP) method [34]. SHAP assigns values that measure the marginal contribution of each variable to the prediction outputs produced by the machine learning model.

### Evaluation of model performance

The XGBoost algorithm was trained using multiple feature subsets obtained through ADT, CVFE and HFE methodologies in conjunction with the training data. Model performance on the test dataset was measured using the metrics defined in Equations 1-4, where TP, TN, FP, and FN represent true positives, true negatives, false positives, and false negatives, respectively. Among the evaluation criteria, accuracy reflects the ratio of correctly predicted cases to the overall number of observations in the dataset, serving as a key indicator of the classifier’s predictive capability.

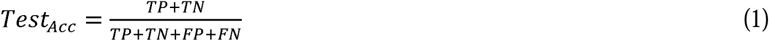

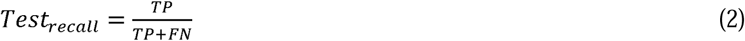

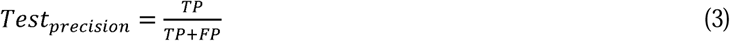

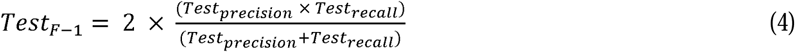

In addition, we evaluated the model using recall and precision metrics. Recall assesses how effectively the model identifies genuine positive instances, whereas precision represents the ratio of correctly predicted positives to the total number of positive predictions made by the model. To provide a balanced measure of both recall and precision, we computed the F1 score, which is the harmonic mean of the two. We also assessed the model’s binary classification performance using the Area Under the Curve (AUC). Higher AUC values indicate stronger discriminative capability, with a score of 1 representing perfect classification and 0.5 corresponding to performance no better than random guessing.

Table 2 presents a comparative assessment of XGBoost model performance using feature subsets derived from ADT, CVFE, and HFE selection methods. Correspondingly, Supplementary Figure S3 (**Supplementary Material**) displays the confusion matrices associated with each reduced feature configuration. Among the evaluated models, the subset generated through CVFE with parameters *c* = 4, *e* = 10, and *p* = 0.6 yielded superior predictive outcomes relative to the ADT- and HFE-based models. Notably, the highest-performing model correctly identified 124 out of 156 individuals diagnosed with OUD.

### Features selected via the CVFE method

The highest-performing model in our analysis was the XGBoost classifier trained using the feature subset identified by the CVFE method, configured with parameters *c* = 4, *e* = 10, and *p* = 0.6. Figure 5 presents a SHAP summary bar chart, which ranks the importance of the selected features based on SHAP value analysis applied to this specific model. From an initial set of 32 candidate variables, the CVFE approach retained 29 features. Among them, the top 10 most influential features—ranked by their SHAP values—were: EDUD5PNRMIS (past-year misuse of pain relievers), PYUD5ALC (alcohol use disorder within the last year), CATAG6 (age group), ASTHMAEVR (history of asthma), SUTRTPY (receipt of substance use treatment in the past year), IREDUHIGHST2 (educational attainment), IRHHSIZ2 (number of household members), INCOME (total household income), IRMARIT (marital status), and NEWRACE2 (race/ethnicity).

**Figure 5.**
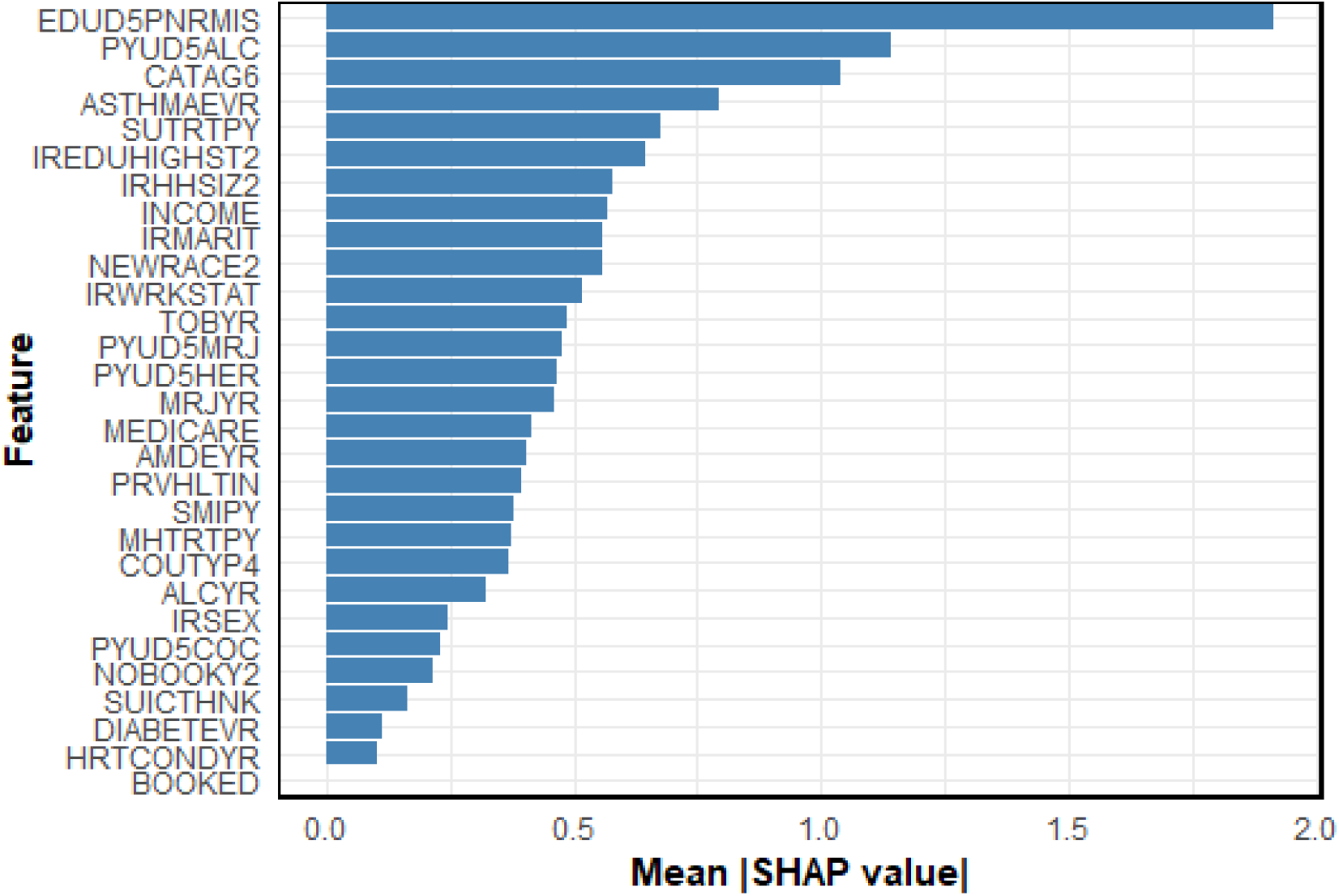
The ranking of feature relevance as identified by the XGBoost framework. The horizontal axis represents the average magnitude of SHAP values for each variable, reflecting their typical influence on prediction outcomes. Features are arranged in decreasing order of predictive contribution.

### Analysis of feature contributions

Figure 6 presents boxplots illustrating the distribution of SHAP values for the categories within the top 10 categorical features identified in Figure 5. Since elevated SHAP values correspond to an increased opioid risk for patients, these plots allow direct interpretation of risk levels across feature categories. For instance, as shown in Figure 6(a), individuals who have never misused pain relievers exhibit a low opioid risk, whereas those classified as “Yes (misused)” show a substantially higher risk. Similarly, Figure 6(b) reveals that patients without an alcohol use disorder have a reduced opioid risk. The SHAP values for age categories, displayed in Figure 6(c), indicate that older individuals carry a higher opioid risk. Surprisingly, Figure 6(d) shows that a history of asthma is linked to a lower opioid risk. Recent substance use treatment correlates with an elevated opioid risk as demonstrated in Figure 6(e). Educational attainment at the college graduate level or above is associated with decreased risk (Figure 6(f)). Finally, factors such as larger household size, lower family income, widowed marital status, and being Non-Hispanic Native American or Alaska Native correspond to increased opioid risk, as shown in Figures 6(g) - 6(j).

**Figure 6.**
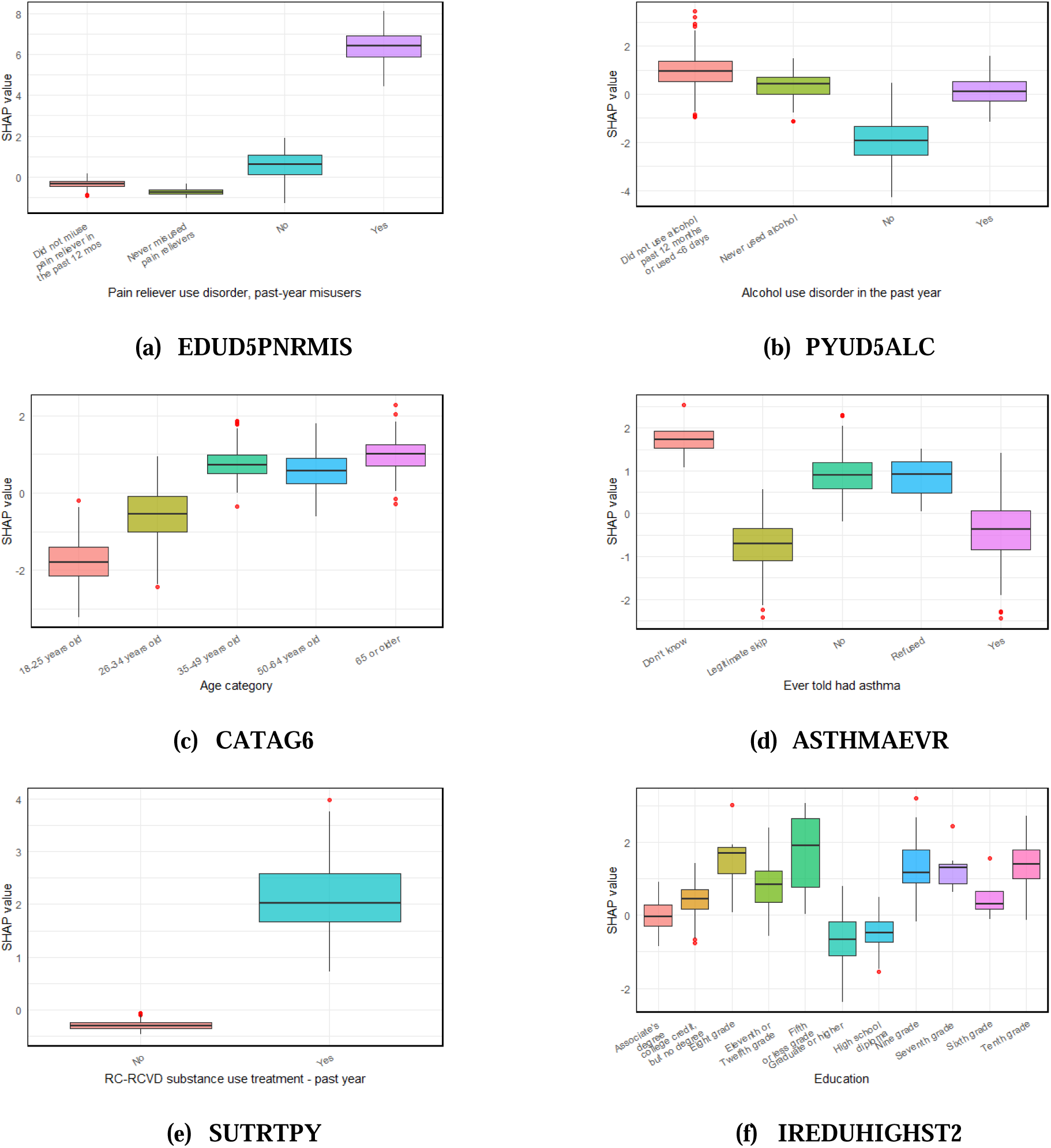
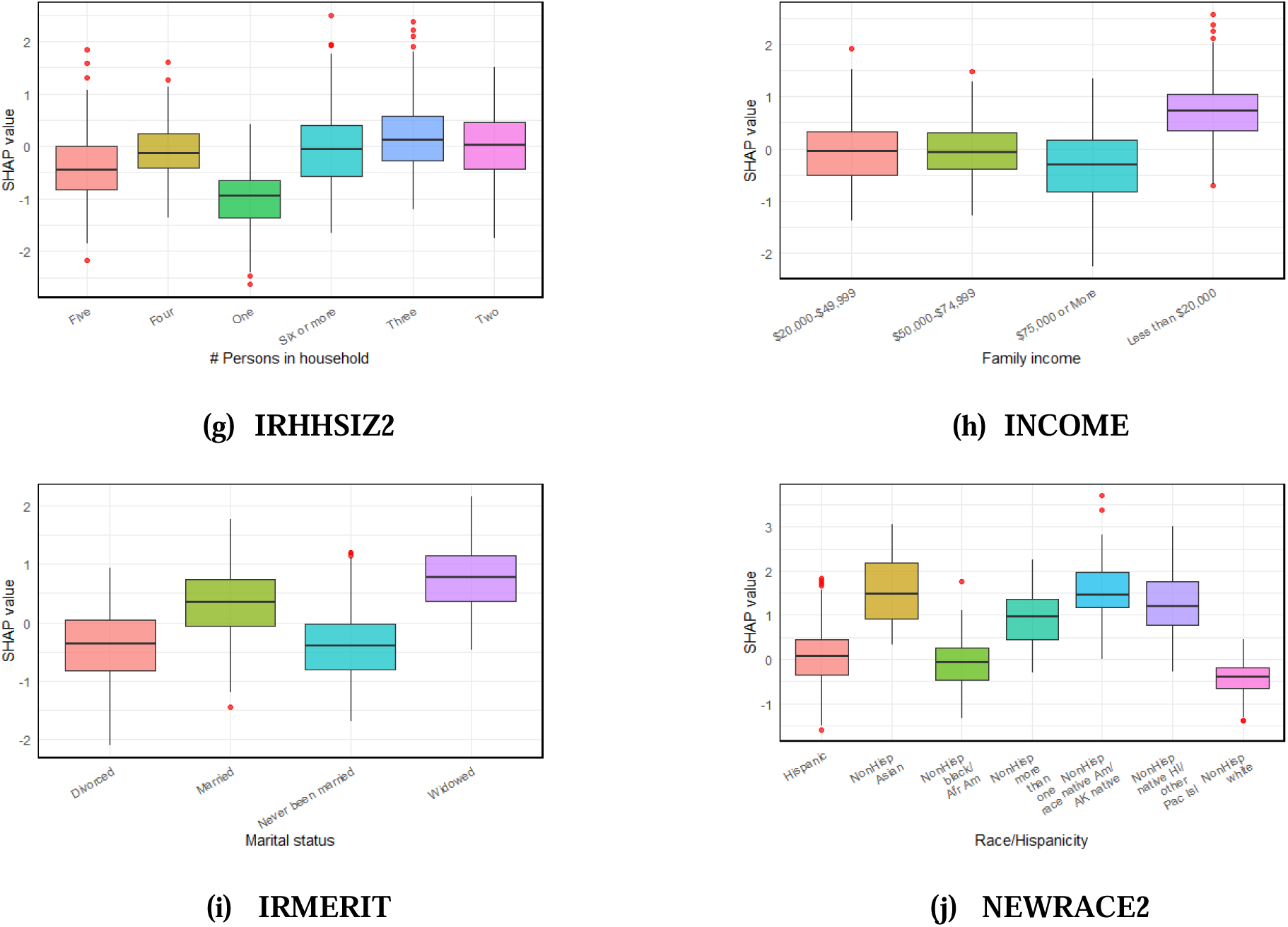
SHAP value distribution for the top 10 categorical features selected by the model.

## Discussion and conclusions

Tailoring predictions to individual patients is critical for enhancing care in those vulnerable to OUD. To meet this goal, we constructed a machine learning prediction framework built on a selectively refined set of features. By removing unnecessary and redundant variables, we improved the model’s clarity and interpretability. Utilizing this streamlined feature set, we assessed the performance of the XGBoost algorithm. Our approach effectively pinpointed important predictors linked to OUD, offering valuable insights for personalized risk evaluation, clinical decision-making, and the development of focused therapeutic strategies. The developed web application integrates the best feature selection method, CVFE, and allows users to apply this approach for predicting OUD risk on new external datasets. Additionally, users have the option to augment the training data with extra samples and perform SHAP-driven analysis to investigate the impact of important features.

Aligning with prior research, our model recognized age and ethnicity as among the most significant predictors. For example, a study utilizing a hierarchical attention transformer model also emphasized these features within the top 10 predictors [3]. However, that analysis lacked a detailed hierarchy of feature importance, which limited its explanatory power. Similarly, Hasan et al. [22] employed several machine learning approaches—including logistic regression, random forest, decision trees, and gradient boosting—on extensive healthcare claims data. Their highest-performing random forest model identified age, heroin poisoning, and drug abuse as key predictors. Our results are consistent with these observations; notably, younger age was linked to a decreased estimated risk for OUD, indicating a potential protective effect in specific scenarios. Additionally, Banks et al. [23] constructed a personalized prediction model for opioid-related outcomes, where age, substance use, and alcohol consumption ranked as the most influential features. The convergence of these findings supports the robustness and clinical relevance of our predictive model.

Importantly, our model highlighted pain reliever misuse as the foremost predictor for OUD. This finding is consistent with prior research underscoring the pivotal influence of prescription opioid misuse in the onset of OUD [35–37]. Recognizing such critical predictor is of great clinical importance, as it can guide early detection efforts, improve risk assessment, and shape targeted intervention plans. Additionally, the other key features identified by our model carry substantial potential to enhance individualized patient management and improve the efficient distribution of healthcare resources. While our results are encouraging, there are important limitations to consider. The models were developed using historical datasets, some of which contained missing data that were addressed through imputation methods [32]. This preprocessing step may have introduced bias, potentially affecting the calculated feature importance values. Furthermore, features with limited representation due to small sample sizes might have disproportionately influenced the findings. Additionally, the current model does not support time-to-event or longitudinal prediction tasks, limiting its application for monitoring the progression of OUD over time. Future work should focus on enhancing the model to accommodate these types of analyses.

In conclusion, we established and validated a machine learning framework to pinpoint critical determinants of OUD using data from the NSDUH survey. While the model shows encouraging predictive capability, there remains room for refinement, especially regarding improvements in AUC and other performance indicators. Future efforts will aim to integrate more detailed, high-dimensional datasets—such as multi-omics profiles and behavioral health metrics—to boost the model’s

accuracy. This research advances the development of personalized strategies for opioid misuse prevention and treatment, ultimately aiming to enhance patient care outcomes.

## Supporting information

The supplementary material for this article is provided herewith the manuscript. (DOCX)

## Authors’ contributions

SA: Conceptualization, data collection, formal analysis, software implementation, validation, visualization, and writing manuscript. JHM: Conceptualization, supervision, reviewing analyses, and editing the manuscript. All authors read and approved the final manuscript.

## Ethical approval statement

The study was conducted in accordance with the Declaration of Helsinki.

## Data Availability

All data produced are available online at https://github.com/suraiya14/OUDP.

