## Supplementary Materials for "Identifying Key Predictive Features for Opioid Use Disorder Using Machine Learning"

### 1 Supplementary Data

#### 1.1 Training dataset

The training dataset can be accessible at https://github.com/suraiya14/OUDP.

#### 1.2 Testing dataset

The testing dataset can be accessible at https://github.com/suraiya14/OUDP.

### 2 Supplementary Tables

#### 2.1 Feature Set before applying feature reduction algorithm

**Table S1.** List of features used for feature reduction analysis

| CATAG6 | IRSEX | NEWRACE2 | IREDUHIGHST2 | INCOME | IRMARIT | IRWRKSTAT | COUTYP4 |
| --- | --- | --- | --- | --- | --- | --- | --- |
| PYUD5ALC | PYUD5MRJ | PYUD5COC | PYUD5HER | EDUD5PNRMIS | TOBYR | ALCYR | MRJYR |
| COCYR | HERYR | AMDEYR | SMIPY | SUICTHNK | IRSUITRYYR | SUTRTPY | MHTRTPY |
| ASTHMAEVR | DIABETEVR | HRTCONDYR | BOOKED | IRHHSIZ2 | NOBOOKY2 | MEDICARE | PRVHLTIN |

#### 2.2 ADT reduced features

**Table S2.** List of features obtained from ADT

| EDUD5PNRMIS | PYUD5ALC | CATAG6 | SUTRTPY | MRJYR |
| --- | --- | --- | --- | --- |
| SMIPY | IREDUHIGHST2 | PYUD5HER | DIABETEVR | IRWRKSTAT |

#### 2.3 CVFE reduced features

In the following tables S3-S7, *c*, *e* and *p* indicate count of disjoint sub-parts, count of iterations and ratios of recurring iterations for the extraction of common features, respectively.

**Table S3.** List of features obtained from CVFE (*c* = 2, *e* = 5, *p* = 0.2)

| EDUD5PNRMIS | SUTRTPY | PYUD5HER | SMIPY | MEDICARE | ASTHMAEVR | PYUD5ALC | CATAG6 |
| --- | --- | --- | --- | --- | --- | --- | --- |
| DIABETEVR | AMDEYR | MHTRTPY | SUICTHNK | MRJYR | PYUD5COC | IRWRKSTAT | PRVHLTIN |
| IREDUHIGHST2 | TOBYR | NEWRACE2 | PYUD5MRJ | ALCYR | INCOME | IRMARIT | NOBOOKY2 |
| COUTYP4 | IRHHSIZ2 | HRTCONDYR | IRSEX | BOOKED | IRSUITRYYR |  | |

**Table S4.** List of features obtained from CVFE (*c* = 3, *e* = 5, *p* = 0.6)

| EDUD5PNRMIS | SUTRTPY | PYUD5HER | SMIPY | AMDEYR | MEDICARE | CATAG6 | PYUD5ALC |
| --- | --- | --- | --- | --- | --- | --- | --- |
| ASTHMAEVR | IRWRKSTAT | DIABETEVR | MHTRTPY | INCOME | PYUD5MRJ | PRVHLTIN | HRTCONDYR |
| MRJYR | IREDUHIGHST2 | TOBYR | PYUD5COC | IRHHSIZ2 | NOBOOKY2 | NEWRACE2 | ALCYR |
| BOOKED | COUTYP4 | IRMARIT | IRSEX | SUICTHNK |  | | |

**Table S5.** List of features obtained from CVFE (*c* = 4, *e* = 10, *p* = 0.6)

| EDUD5PNRMIS | SUTRTPY | MEDICARE | ASTHMAEVR | PYUD5HER | SMIPY | PYUD5ALC | CATAG6 |
| --- | --- | --- | --- | --- | --- | --- | --- |
| DIABETEVR | AMDEYR | MHTRTPY | MRJYR | SUICTHNK | IRWRKSTAT | PYUD5COC | PRVHLTIN |
| IREDUHIGHST2 | TOBYR | NEWRACE2 | PYUD5MRJ | ALCYR | INCOME | IRMARIT | NOBOOKY2 |
| COUTYP4 | IRHHSIZ2 | HRTCONDYR | IRSEX | BOOKED |  |  |  |

**Table S6.** List of features obtained from CVFE (*c* = 8, *e* = 5, *p* = 0.6)

| EDUD5PNRMIS | CATAG6 | PYUD5ALC | SUTRTPY | ASTHMAEVR | IRWRKSTAT | MHTRTPY | INCOME |
| --- | --- | --- | --- | --- | --- | --- | --- |
| PYUD5MRJ | PRVHLTIN | MEDICARE | IREDUHIGHST2 | MRJYR | IRHHSIZ2 | TOBYR | NEWRACE2 |
| ALCYR | PYUD5COC | COUTYP4 | IRMARIT | IRSEX |  |  |  |

**Table S7.** List of features obtained from CVFE (*c* = 10, *e* = 10, *p* = 0.2)

| EDUD5PNRMIS | CATAG6 | PYUD5ALC | ASTHMAEVR | IRWRKSTAT | INCOME | MHTRTPY | PYUD5MRJ |
| --- | --- | --- | --- | --- | --- | --- | --- |
| SUTRTPY | PRVHLTIN | IREDUHIGHST2 | TOBYR | IRHHSIZ2 | MEDICARE | NEWRACE2 | MRJYR |
| ALCYR | COUTYP4 | IRMARIT | AMDEYR | IRSEX |  |  |  |

#### 2.4 HFE reduced features

The lists of features obtained from the HFE method are presented in Tables S8–S10. In each table, the first 8, 16, and 24 features correspond to β values of 25, 50, and 75, respectively.

**Table S8.** List of features obtained from HFE with *β* = 25

| EDUD5PNRMIS | PYUD5COC | SUTRTPY | TOBYR |
| --- | --- | --- | --- |
| PYUD5ALC | PYUD5HER | BOOKED | NOBOOKY2 |

**Table S9.** List of features obtained from HFE with *β* = 50.

| EDUD5PNRMIS | PYUD5COC | SUTRTPY | TOBYR | PYUD5ALC | PYUD5HER | BOOKED | NOBOOKY2 |
| --- | --- | --- | --- | --- | --- | --- | --- |
| PRVHLTIN | MHTRTPY | IRWRKSTAT | MRJYR | PYUD5MRJ | SMIPY | AMDEYR | DIABETEVR |

**Table S10.** List of features obtained from HFE with *β* = 75.

| EDUD5PNRMIS | PYUD5COC | SUTRTPY | TOBYR | PYUD5ALC | PYUD5HER | BOOKED | NOBOOKY2 |
| --- | --- | --- | --- | --- | --- | --- | --- |
| PRVHLTIN | MHTRTPY | IRWRKSTAT | MRJYR | PYUD5MRJ | SMIPY | AMDEYR | DIABETEVR |
| ASTHMAEVR | INCOME | IREDUHIGHST2 | CATAG6 | SUICTHNK | IRMARIT | HERYR | COCYR |

**3. Supplementary Figures**

**3.1 ADT summary plot**

**Figure S1 –** A alternative decision tree representation using the reduced ADT feature set.

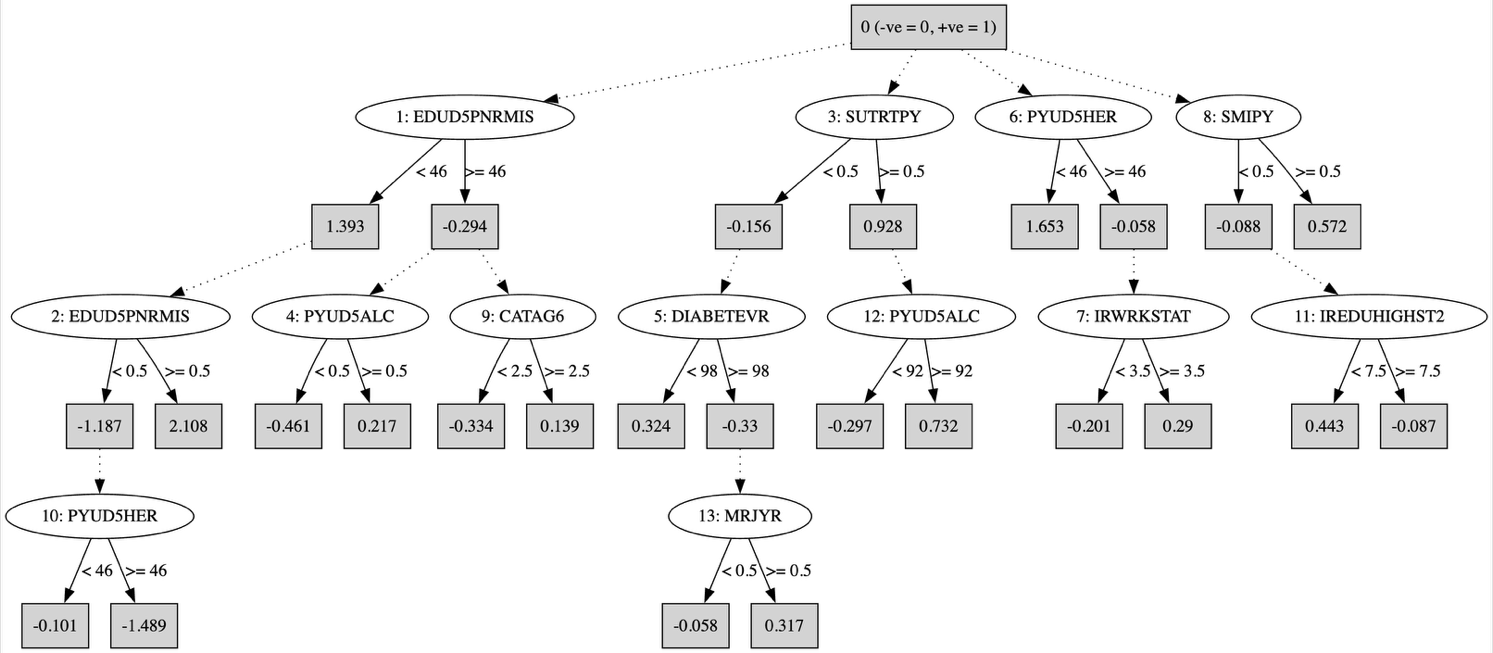

**3.2 Hypergraph**

**Figure S2 –** Hypergraph representation of the training dataset

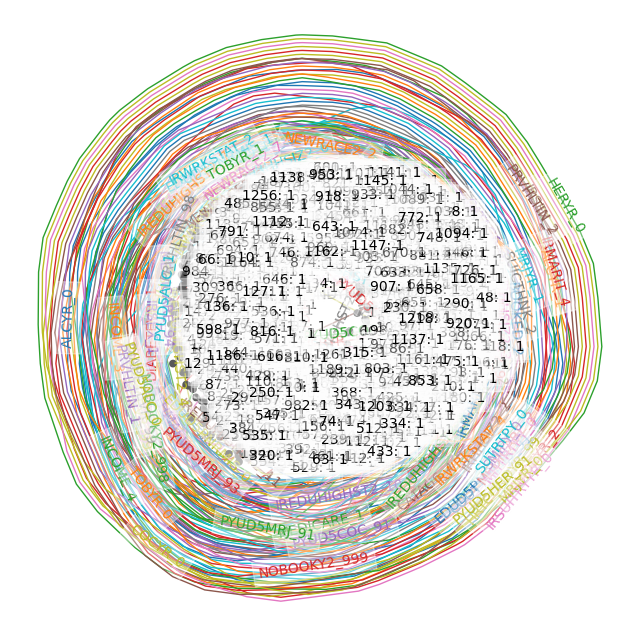

**3.3 Confusion matrices**

**Figure S3 –** Confusion matrices of the machine learning model built with the reduced feature sets

| 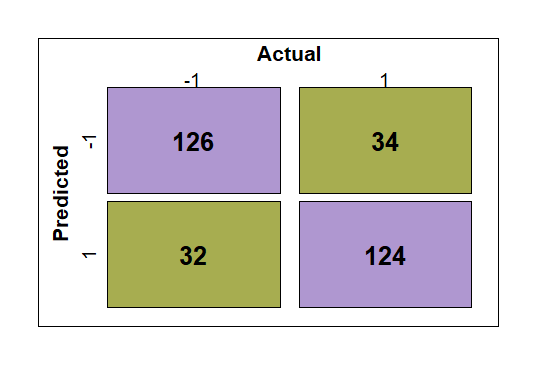   (a) **CVFE (*c* = 2, *e* = 5, *p* = 0.2)** | 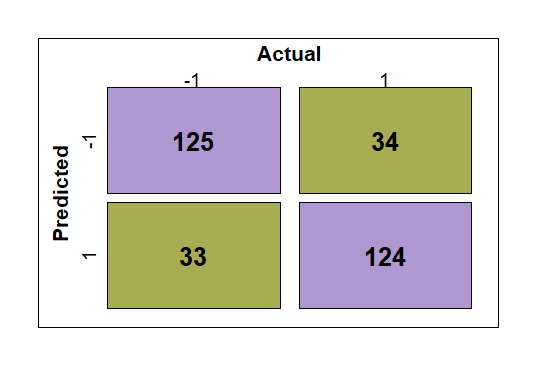   (b) **CVFE (*c* = 3, *e* = 5, *p* = 0.6)** | 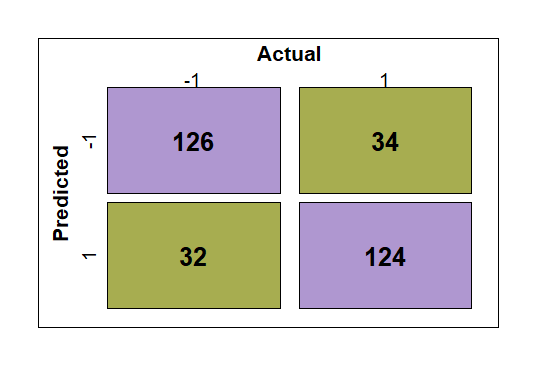   (c) **CVFE (*c* = 4, *e* = 10, *p* = 0.6)** |
| --- | --- | --- |
| 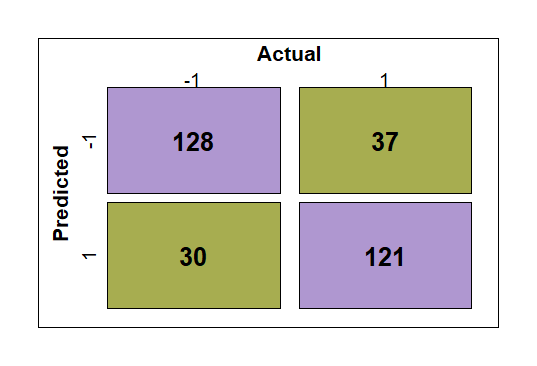   (d) **CVFE (*c* = 8, *e* = 5, *p* = 0.6)** | 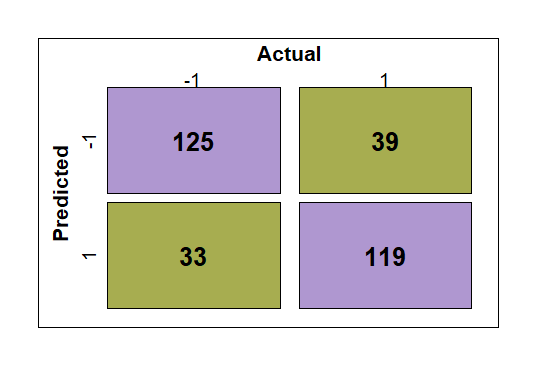   (e) **CVFE (*c* = 10, *e* = 10, p=0.2)** | 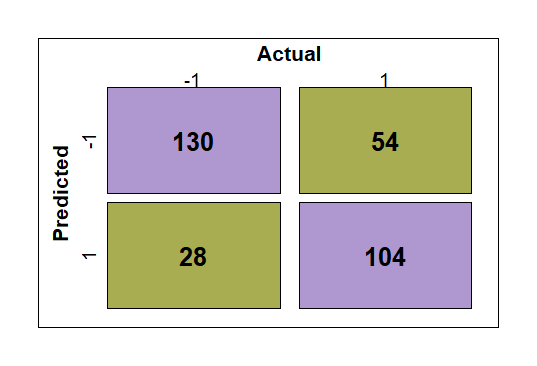   (f) **HFE (***β* **=25)** |
| 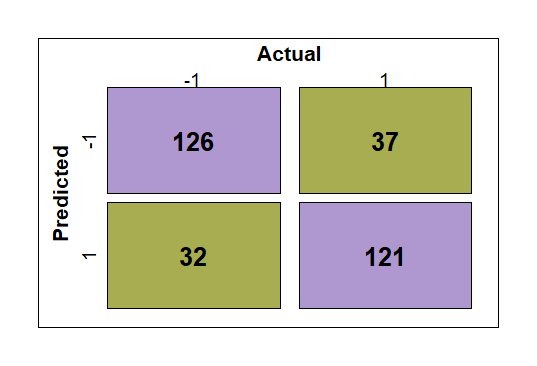   (g) **HFE (***β* **= 50)** | 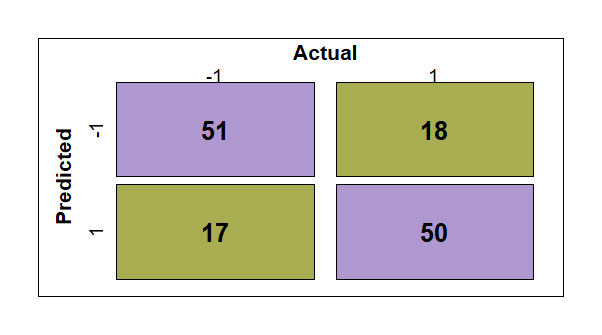   (h) **HFE (***β* **= 75)** | 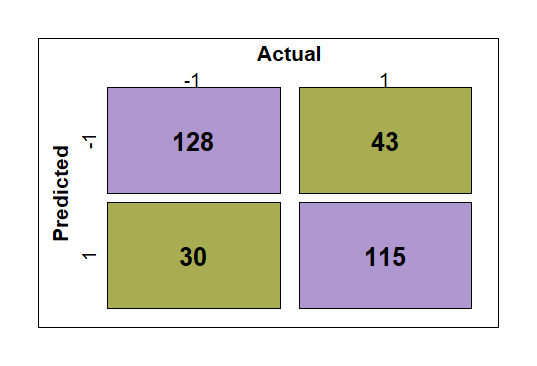   (i) **ADT** |
